# PCGS: biomarker and risk group identification for Pediatric Cancers via explainable Graph neural networks with Shapley values

**DOI:** 10.64898/2026.08.27.26361540

**Authors:** Zanyu Shi, Aishwarya Budhkar, Waqas Amin, Karen E. Pollok, Jing Su, Kun Huang

## Abstract

Improvements in data availability, sharing, and integration, together with the development of explainable artificial intelligence (XAI) techniques, are advancing precision medicine for pediatric cancer by facilitating diagnosis, biomarker discovery, and drug development. Data sharing commons and initiatives like the Childhood Cancer Data Initiative (CCDI) provide access to pediatric-specific genomic and clinical data cohorts and improve data availability for pediatric cancer research. Based on CCDI, a scalable AI platform, Graph Artificial Intelligence for Pediatric Oncology (GAIPO), integrates various data modalities from bulk and single-cell omics data to clinical information. Such multi-modal data facilitates the training and development of advanced XAI models for pediatric cancers. We then developed an end-to-end multi-modality framework, PCGS, for pediatric cancer by incorporating omics-specific representation learning via GNN models with cross-attention fusion and multi-objective learning for downstream tasks such as classification, clustering, and survival analysis. This framework outperforms previous supervised multi-omics integration baseline approaches based on glioma and Wilms tumor cohorts and enables GNN model explainability via Shapley value-based feature attribution approaches to explain the contributions of gene-level features across various biomedical tasks, including classification and survival. Given specific background samples (e.g., age groups, sex, grades) as baselines, this explainable GNN model estimates and ranks the importance scores for input features from each omics modality. It identifies background-specific key features for biomarker discovery, risk group identification, and survival analysis in glioma and Wilms tumor, with potential applicability to other pediatric cancers.

## 1. Introduction

Advances in artificial intelligence (AI) are transforming precision medicine for pediatric cancers, enabling progress in diagnosis and prognosis through biomarker discovery, drug development, and survival analysis^1,2^. Graph Neural Networks (GNNs) offer distinct advantages in flexibly integrating heterogeneous, multimodal data from diverse sources^3,4^, showcase robust representation learning capabilities for both structural graphs (e.g., single-cell spatial transcriptomics^5^) and non-structural graphs (e.g., sample similarity networks^6^), accelerating drug and biomarker discovery in cancer research. Moreover, explainable AI (XAI) models are widely applied to multi-omics integrative analysis using sample- or gene-similarity networks to facilitate biomarker discovery predominantly in adult cancer research^7^ and increasingly extended to pediatric cancers^8^.

However, challenges remain in pediatric cancer research, including the rarity of pediatric cancer samples, limited data availability^1,9^, intrinsic heterogeneity of pediatric tumors^10^, diverse clinical presentations and treatment responses compared to adult cancers^11^, and ethical issues related to data privacy and algorithmic transparency^12^. First, there are generally larger and more accessible datasets for adult cancers, for example, The Cancer Genome Atlas (TCGA)^13^. Such datasets facilitate the model training process and enable the development of robust models by providing extensive clinical information, molecular biomarkers and genomic data. In contrast, pediatric cancers are relatively rare diseases and thus less data are available^9^. This scarcity of data can hinder the development of highly accurate models and new model frameworks. Moreover, there exists inherent complexity in pediatric oncology involving unique genetic and environmental interactions, which are known to be different from adult malignancies in terms of cellular origins, epidemiology, genetic intricacies, key driver mutations, and fundamental mutational processes^10^. This complexity requires not only more specialized datasets but also sophisticated modeling techniques that can handle such diverse and less structured data. Additionally, pediatric cancers often exhibit different clinical representations and responses to treatment compared to adult cancers, requiring models specifically trained on pediatric data to achieve similar levels of accuracy and applicability^11^.

Many recent efforts have aimed to create a collaborative network centered on childhood cancer care and research to address the above challenges. For example, data sharing commons and initiatives such as the Childhood Cancer Data Initiative (CCDI)^14^, Gabriella Miller Kids First Data Resource Portal (PedcBioPortal)^15^ and The cBioPortal for Cancer Genomics^16^, aim to improve data availability for pediatric cancer research by providing access to genomic and clinical data specifically from pediatric cohorts. CCDI seeks to deepen the understanding of pediatric cancer biology and foster improvements in the diagnosis, treatment, and long-term survival outcomes for pediatric cancer patients^17^. To streamline the use of the CCDI resources such as multimodal clinicogenomics and clinicospatial data for AI model training and implementation, a generic graph AI platform, Graph Artificial Intelligence for Pediatric Oncology (GAIPO), as well as the standards of data, model, and pipeline, were developed. Leveraging the GAIPO standards and workflow, researchers can access and utilize multimodal data to train specific deep learning models for pediatric cancers. Furthermore, with the development of data-sharing schemes and infrastructure, biomarker and drug discovery and risk group identification via multi-omics analysis for pediatric cancer diagnosis and prognosis has also garnered significant interest^18,19^. Utilizing multi-omics data to develop end-to-end GNN models for multi-objective tasks such as classification and survival analysis also enables downstream tasks like stratifying patients into risk groups^6^. This can help clinicians conduct preventive interventions, improve survival outcomes, and tailor suitable personalized treatment strategies for pediatric cancer patients^1,19^.

Approaches for explainable GNNs in biomarker discovery for human diseases align with the applications of XAIs like post-hoc explainability techniques ^7^. Feature attributions based on the Shapley value are popular for explaining machine learning and deep learning models^20^. Braytee et al. (2024) incorporate Shapley values to provide the interpretability of the predictions via the autoencoder framework that captures non-linear data representations and utilizes tensor analysis for feature extraction^21^. Moreover, DeepSHAP^22^ was designed to produce biased but computationally efficient estimates of marginal Shapley values and provide a rescale rule to propagate attributions through the pipelines of deep neural networks like GNNs with fusion layers.

Such estimated Shapley values can be used to explain the contribution of features like gene-level importance for representation learning, prediction and classification tasks, given specific background samples as baselines^22^.

In this paper, we introduce PCGS (i.e., biomarker and risk group identification for Pediatric Cancers via GNNs with Shapley-value-based explainability), an explainable GNN framework for biomarker discovery and risk-group identification in pediatric cancers using Shapley values. Unlike most explainable GNN applications that have been primarily developed for adult cancers, PCGS leverages GNN layers with cross-attention fusion layers to integrate pediatric multi-omics data with clinical data and incorporates Shapley value-based feature attribution to address challenges in pediatric oncology. PCGS has the following major contributions:

1. It is a novel explainable GNN framework with an end-to-end pipeline incorporating GNN and fusion layers with multi-objective learning for downstream tasks to integrate multi-omics and clinical data for classification, clustering, and survival analysis.
2. It incorporates Shapley value-based post-hoc feature attribution to rank and identify vital biomarkers in each modality and assess background-specific importance scores for biomarker discovery and risk stratification.
3. Its effectiveness is demonstrated by systematic evaluation on glioma and Wilms tumor datasets with improved predictive performance and interpretable biomarker identification comparing with existing multi-omics integration approaches, which can be adopted for other pediatric precision oncology research.

## 2. Method

In this work, we developed an end-to-end multi-modality framework that employs GNNs to learn representations for each omics modality, integrates cross-attention fusion layers with multi-objective learning for downstream tasks such as classification and survival analysis, and applies Shapley value-based feature attribution to identify key biomarkers for patient stratification, with sensitivity to baseline background selection (Figure 1).

**Figure 1.**
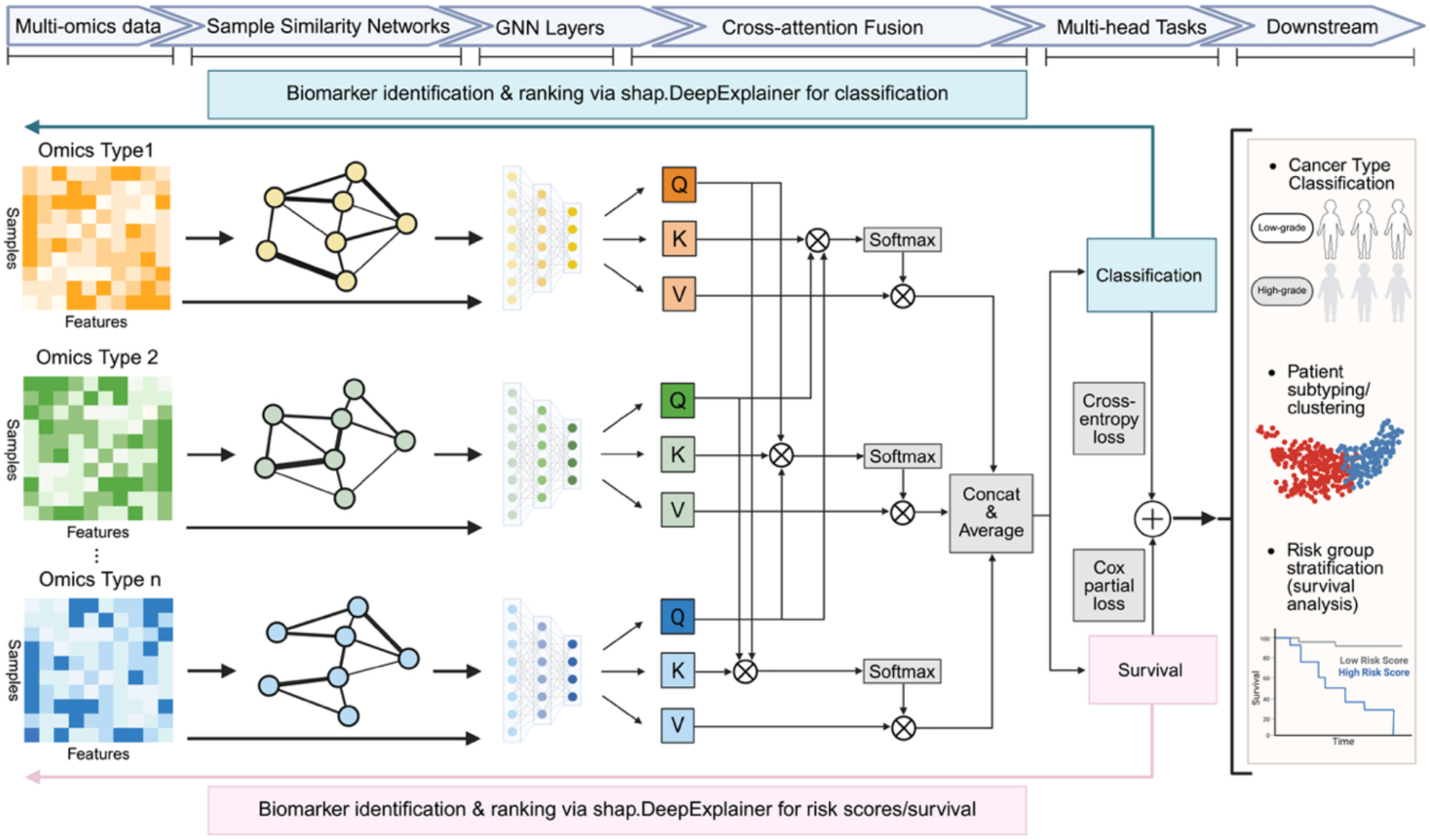
Model framework illustration of PCGS. It provides an end-to-end multi-modality framework for pediatric cancer by incorporating omics-specific representation learning via GNN models with cross-attention fusion and multi-objective learning for downstream tasks such as classification, clustering, and survival analysis. This framework also enables GNN model explainability via Shapley value-based feature attribution approaches to explain the contributions of gene-level features across various biomedical tasks, including classification and risk group identification.

### 2.1. Data preprocessing

In this project we demonstrate PCGS using two cohorts from two pediatric cancers, glioma and Wilms tumor (Table 1). Multi-omics data of glioma containing mRNA sequencing and Copy-number alterations (CNAs) are downloaded from the Pediatric Brain Tumor Atlas (PBTA) ^23^, and Wilms tumor data are downloaded from TARGET: Kidney, Wilms tumor ^24^, including mRNA, methylation, CNA, and microRNA (miRNA) via the cBioPortal ^25^ API. The main classification levels for glioma are high-grade vs. low-grade, and the ones for Wilms tumor are favorable histology Wilms tumor (FHWT) vs. diffuse anaplastic Wilms tumor (DAWT). The raw numbers of features for each dataset and each omics data type are listed in Table 1.

**Table 1.** Summary of pediatric cancer datasets.

| Cancer | Categories | # raw features | # features after pre-selection |
| --- | --- | --- | --- |
| Glioma | 393 patients: | mRNA: 60372 | mRNA: 1000 |
|  | 322 Low-grade<br>71 high-grade | CNA: 39584 | CNA: 1000 |
| Wilms tumor | 120 patients: | mRNA: 20430 | mRNA: 1000 |
|  | 82 FHWT | microRNA: 1872 | microRNA: 500 |
|  | 38 DAWT | Methylation: 15020 | Methylation: 500 |

Proper data preprocessing for omics data is carried out to reduce human and experimental errors and improves model performance and explainability. We first filtered out features with no signal (i.e., zero mean) or low variance. Different variance filtering thresholds were set for various omics data types across different cancers (i.e., 0.1 for mRNA expression data, 0.05 for methylation data, and 0.1 for microRNA data). Moreover, we further preselected omics features using statistical tests since redundant features in omics data may negatively influence model performance. ANOVA *F*-value with false-discovery rate (FDR) correction by the Benjamini-Hochberg procedure for multiple-testing compensation was applied to evaluate whether a feature was significantly different across levels in a classification task. Additionally, to avoid redundant and highly correlated features and preserve complementary information, we conduct principal component analysis (PCA) and choose preselected features per omics type so that its first principal component explains less than 50% of the variance. The numbers of preselected features for each omics data are also shown in Table 1. Finally, we scaled each type of omics data to the range of (0, 1) respectively via linear transformations.

### 2.2. GNN Model Framework

In general definitions, a graph can be defined as *G* = (*V, ε*) with adjacency matrix *A* ∈ ℝ^*N*×*N*^, where |*V*| = *N* is the number of nodes in the graph and |*ε*| = *N*^*e*^ is the number of edges. The hidden state ***h***_*v*_ and the output vector ***o***_*v*_ of a node *v* can be further defined for graph representation learning. We viewed a patient with relevant features for each omics data as a node *v* ∈ *V* characterized in a graph *G* = (*V, ε*), with edges *e* ∈ *ε* representing the relationships between nodes based on the sample similarity network generated by cosine similarity. We employed GNN layers that can be selected from Graph Convolution Networks (GCN) ^26^, Graph Isomorphism Networks (GIN) ^27^, and Graph Attention Networks (GAT) ^28^ to learn latent representations for each omics type by utilizing both the features of each node and their relationships (i.e., edges). We then applied fusion methods like cross-attention fusion and simple fusion layers to integrate such latent-space information into multi-objective learning tasks with branches of classification and survival analysis. Specifically, cross-attention-based multimodal representation simultaneously extracts features from various omics^29^, while simple fusion was followed by CANDECOMP/PARAFAC (CP) decomposition ^30^ using regularized ALS (RALS) algorithms ^21^. Given a Query matrix 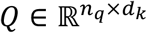 from modality A (target modality), a Key matrix 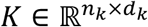 and a Value matrix 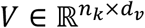 from modality B (source modality), and *d*_*k*_ representing dimensionality of keys/queries used for scaling, the cross-attention output is computed as

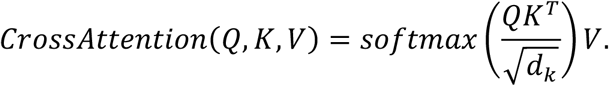

The attention weights, *softmax*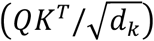, represent how strongly each query attends to the source elements, and the weighted sum of *V* captures the context from modality B relevant to modality A^31^. Moreover, classification with cross-entropy loss and survival with negative Cox partial log-likelihood loss were defined and added with weights as the branches of multi-objective learning tasks for end-to-end model training. Model classification performance is evaluated by accuracy and F1 scores, and survival or risk score prediction is evaluated using the concordance index (C-index), a metric that measures a model’s ability to correctly rank the order of events for survival analysis, such as time to death, based on predicted risk scores ^32^. 5-fold cross-validation was applied for fine-tuning hyperparameters and comparing model performance with baseline models. Furthermore, PCGS can utilize the output from cross-attention fused layers or components derived from tensor decomposition for downstream tasks such as classification, clustering, and survival analysis. For example, it calculates risk scores for patient subtyping via hierarchical clustering or risk split by median and conducts survival analysis for various pediatric cancers, evaluated by Kaplan-Meier plots and log-rank test p-values. We then performed hierarchical clustering and risk group split by median (high vs. low) on the learned latent representation and evaluated whether the resulting patient subtypes also show distinct survival patterns.

### 2.3. Feature attribution and model explainability

Post-hoc Shapley value-based feature attribution method was applied to estimate local feature contribution and identify important biomarkers in each modality. The process ranked and selected the top biomarkers contributing to classification or survival for baseline samples from specific backgrounds. Because exact computation is infeasible for high-dimensional deep learning models, for PCGS, we implemented the model-agnostic approximation method of marginal Shapley values in DeepSHAP ^22^ that combines interventional SHAP semantics with efficient backpropagation-based attribution rules. This allows attribution scores to be propagated through the differentiable components of the PCGS architecture including GNNs with cross-attention fusion and multi-head task layers. Specifically, it recursively passes the multipliers of approximation of Shapley values backwards through the neural networks ^33^ from the output layers of two branches of multi-head tasks (i.e., classification and survival) to the input GNN layers for representation learning in each omics type. In addition, randomly selected or preselected samples with a specific background as baselines are allowed to compute Shapley values of input features tracing back from the output layer of multi-head tasks (i.e., classification and survival) to the input layers of omics-specific GNNs and conduct explanations with different baselines. For example, we can identify the features that are most important for survival compared to a baseline sample selected from the general population of patients with low-grade and high-grade glioma, or a baseline sample that includes only high-grade glioma patients. The contribution of biomarkers in each omics modality for such specific samples and tasks is evaluated and ranked based on estimated Shapley values. For instance, the top genes that significantly impact the survival of patients with high-grade glioma can be identified.

For PCGS, Shapley value-based attribution is used to quantify how individual input features contribute to a patient-specific prediction in pediatric cancer tasks such as subtype classification and survival modeling. In the Shapley framework, each feature is treated as a player in a coalition game, and the attribution score for feature *i*measures its average marginal contribution across all subsets of the remaining features. Let *M* = (1, *…, m*) denote the full set of input features and let *v*(*S*) be a set function that maps a coalition *S* ⊆ *M* to a scalar model value. The Shapley value for feature *i*is defined as

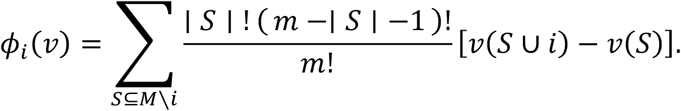

Equivalently, this quantity can be interpreted as the average marginal contribution of feature *i* over all possible feature orderings. For a deep model *f*(*x*): ℝ^*m*^ → ℝ, exact computation of Shapley values is generally intractable, especially in multimodal graph neural network pipelines.

Given a deep learning model *f*(⋅): ℝ^*m*^ → ℝ, an explicand sample *x*^*e*^ ∈ ℝ^*m*^, and a baseline or background cohort *D*_0_, Shapley value-based local attribution can be defined under an interventional setting relative to a baseline sample *x*^*b*^ or a baseline cohort *D*_*b*_. For a subset *S* ⊆*M*, the coalition value is evaluated using a spliced sample χ^*S*^, which is constructed by taking feature values from the explained patient for coordinates in *S*and replacing the remaining coordinates using baseline values:

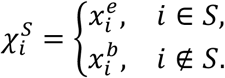

where *x*^*b*^ is sampled from the empirical baseline distribution *D*_0_. The corresponding interventional conditional expectation is

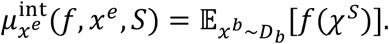

The interventional Shapley value of feature *i* for patient *x*^*e*^ relative to background cohort *D*_0_ is

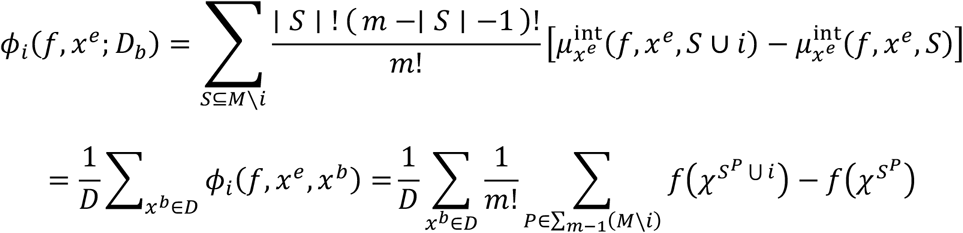

where *D*_*b*_ denotes the empirical distribution of the baseline dataset and assigns equal probability to each sample in the selected baseline dataset. This formulation allows feature importance to be interpreted relative to a biologically meaningful reference cohort, such as a mixed glioma population or only high-grade glioma patients, thereby enabling different patient-level biological questions to be addressed.

To apply this attribution framework in PCGS, we explicitly model the multi-omics GNN pipeline. For modality *j*, let *X*_*j*_ denote the modality-specific node-feature matrix and let *A*_*j*_ denote the corresponding patient similarity graph (or adjacency matrix), which is constructed before model training and held fixed during explanation. The omics-specific GNN encoder generates a latent representation

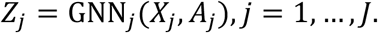

These modality-specific latent embeddings are then integrated by a fusion module:

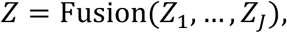

and a downstream task head produces the target output

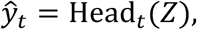

where *t*indexes the prediction task, such as subtype classification or survival risk estimation. Generally, if the entire differentiable PCGS architecture is written as a composition of layer-wise functions 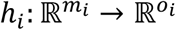, then the full model can be expressed as

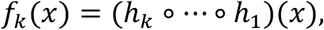

with intermediate mappings

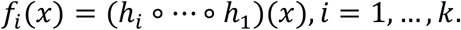

Under this formulation, approximate SHAP values are traced backward from the selected output layer through the task head, fusion module, and omics-specific GNN encoders to the original input features. Following DeepSHAP-style rescale propagation, the layer-wise attribution for modality *j*can be written schematically as

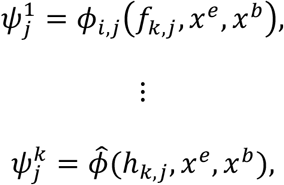

and recursively for intermediate layers,

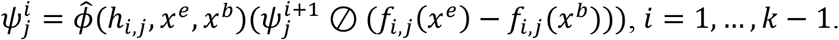

In this expression, 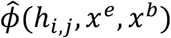 denotes the approximate contribution assigned to layer *h*_*i,j*_ under the DeepSHAP rescale rule, and 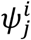 denotes the propagated attribution score at layer *i* for modality *j*. This recursive propagation provides a computationally efficient approximation for decomposing the model output into modality-specific input contributions. Because the patient similarity graph *A*_*j*_ is fixed during explanation, the resulting attribution scores should be interpreted primarily as the contribution of node features conditional on the observed graph structure, rather than as attributions to graph rewiring or edge perturbations.

Finally, PCGS uses these attribution scores for biomarker prioritization. For patient *n*, task *t*, and feature *r*in modality *j*, the patient-level importance score is given by the magnitude of its attribution value,

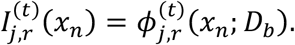

These scores can be ranked within each modality for local explanation of an individual patient. To obtain cohort-level candidate biomarkers, the attribution magnitudes are aggregated across patients:

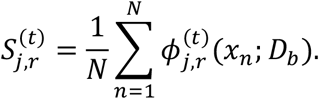

Features with the largest values of 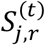 are then prioritized as candidate biomarkers associated with subtype discrimination or survival risk. In this way, PCGS links multimodal graph representation learning with patient-level explainability and cohort-level biomarker discovery in pediatric oncology.

## 3. Results

We compared the model performance for various settings of GNNs for representation learning and fusion layers with baseline methods (Table 2) using glioma and Wilms tumor cohorts. The metrics calculated and averaged based on 5-fold cross-validation suggest that models with GNN layers outperform supervised multi-omics integration baseline approaches including a previously published graph AI model, MOGONET ^6^ across classification (accuracy and F1 scores) and survival analysis (C-index). The combination of GAT and cross-attention achieves the highest accuracy (0.9263 ± 0.0257) and C-index (0.9061 ± 0.0163), and the highest F1 score (0.8334±0.0234) belongs to GIN and cross-attention for glioma, while the combination of GCN and cross-attention gets the highest accuracy (0.9417±0.0333), GIN and simple layer has the highest F1 score (0.9078 ± 0.0684), and GAT and simple layer has the highest C-index (0.6103 ± 0.0486) for Wilms tumor. The results suggest that incorporating omics-specific representation learning via GNNs and cross-attention fusion improves the model performance for classification and survival in pediatric cancer research.

**Table 2.**
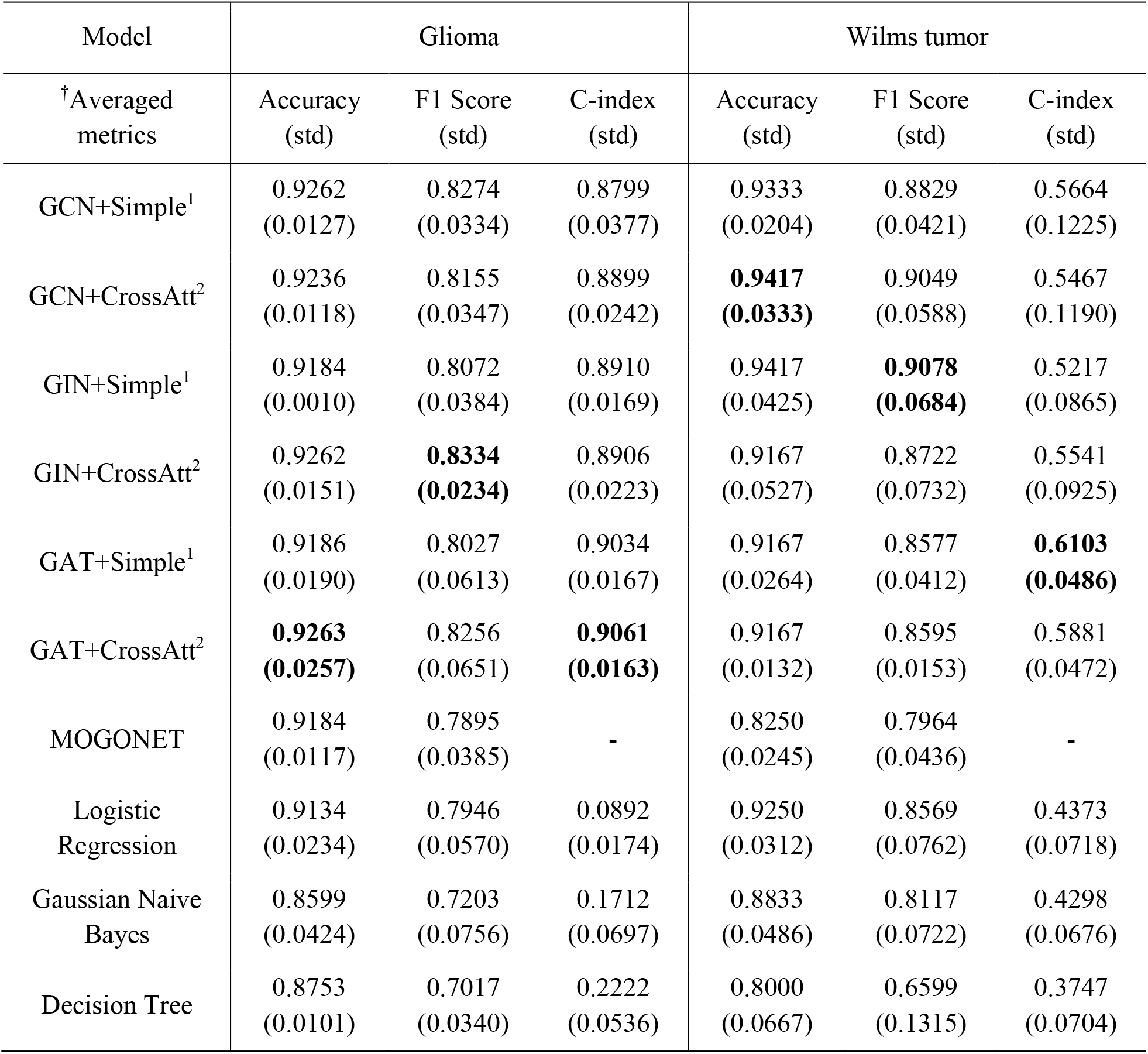
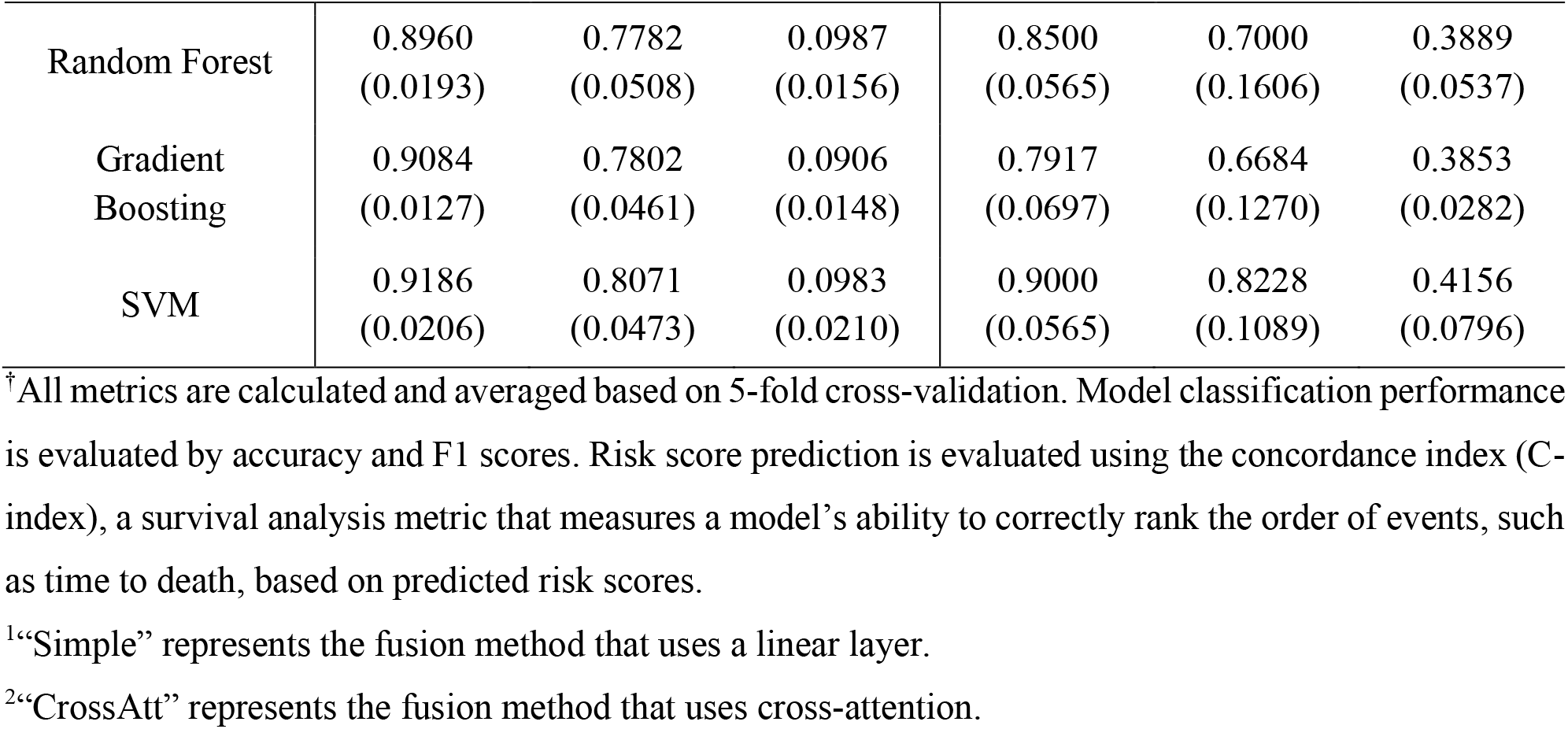
GNNs and baseline model performance comparison via 5-fold cross-validation.

| Model | Glioma |  |  | Wilms tumor |  |  |
| --- | --- | --- | --- | --- | --- | --- |
| <sup>†</sup>Averaged metrics | Accuracy (std) | F1 Score (std) | C-index (std) | Accuracy (std) | F1 Score (std) | C-index (std) |
| GCN+Simple<sup>1</sup> | 0.9262<br>(0.0127) | 0.8274<br>(0.0334) | 0.8799<br>(0.0377) | 0.9333<br>(0.0204) | 0.8829<br>(0.0421) | 0.5664<br>(0.1225) |
| GCN+CrossAtt<sup>2</sup> | 0.9236<br>(0.0118) | 0.8155<br>(0.0347) | 0.8899<br>(0.0242) | **0.9417**<br>**(0.0333)** | 0.9049<br>(0.0588) | 0.5467<br>(0.1190) |
| GIN+Simple<sup>1</sup> | 0.9184<br>(0.0010) | 0.8072<br>(0.0384) | 0.8910<br>(0.0169) | 0.9417<br>(0.0425) | **0.9078**<br>**(0.0684)** | 0.5217<br>(0.0865) |
| GIN+CrossAtt<sup>2</sup> | 0.9262<br>(0.0151) | **0.8334**<br>**(0.0234)** | 0.8906<br>(0.0223) | 0.9167<br>(0.0527) | 0.8722<br>(0.0732) | 0.5541<br>(0.0925) |
| GAT+Simple<sup>1</sup> | 0.9186<br>(0.0190) | 0.8027<br>(0.0613) | 0.9034<br>(0.0167) | 0.9167<br>(0.0264) | 0.8577<br>(0.0412) | **0.6103**<br>**(0.0486)** |
| GAT+CrossAtt<sup>2</sup> | **0.9263**<br>**(0.0257)** | 0.8256<br>(0.0651) | **0.9061**<br>**(0.0163)** | 0.9167<br>(0.0132) | 0.8595<br>(0.0153) | 0.5881<br>(0.0472) |
| MOGONET | 0.9184<br>(0.0117) | 0.7895<br>(0.0385) | - | 0.8250<br>(0.0245) | 0.7964<br>(0.0436) | - |
| Logistic Regression | 0.9134<br>(0.0234) | 0.7946<br>(0.0570) | 0.0892<br>(0.0174) | 0.9250<br>(0.0312) | 0.8569<br>(0.0762) | 0.4373<br>(0.0718) |
| Gaussian Naive Bayes | 0.8599<br>(0.0424) | 0.7203<br>(0.0756) | 0.1712<br>(0.0697) | 0.8833<br>(0.0486) | 0.8117<br>(0.0722) | 0.4298<br>(0.0676) |
| Decision Tree | 0.8753<br>(0.0101) | 0.7017<br>(0.0340) | 0.2222<br>(0.0536) | 0.8000<br>(0.0667) | 0.6599<br>(0.1315) | 0.3747<br>(0.0704) |
| Random Forest | 0.8960<br>(0.0193) | 0.7782<br>(0.0508) | 0.0987<br>(0.0156) | 0.8500<br>(0.0565) | 0.7000<br>(0.1606) | 0.3889<br>(0.0537) |
| Gradient Boosting | 0.9084<br>(0.0127) | 0.7802<br>(0.0461) | 0.0906<br>(0.0148) | 0.7917<br>(0.0697) | 0.6684<br>(0.1270) | 0.3853<br>(0.0282) |
| SVM | 0.9186<br>(0.0206) | 0.8071<br>(0.0473) | 0.0983<br>(0.0210) | 0.9000<br>(0.0565) | 0.8228<br>(0.1089) | 0.4156<br>(0.0796) |

<sup>†</sup>All metrics are calculated and averaged based on 5-fold cross-validation. Model classification performance is evaluated by accuracy and F1 scores. Risk score prediction is evaluated using the concordance index (C-index), a survival analysis metric that measures a model’s ability to correctly rank the order of events, such as time to death, based on predicted risk scores.
<sup>1</sup>“Simple” represents the fusion method that uses a linear layer.
<sup>2</sup>“CrossAtt” represents the fusion method that uses cross-attention.

To further evaluate the model’s performance in downstream tasks, we conducted a survival analysis for patient subtyping using the test sets of glioma and Wilms tumor, based on the output of the GAT with cross-attention fusion and hierarchical clustering (Figure 2). For glioma, the Kaplan-Meier curves of patient subtypes (Figure 2.a) were split by cancer type (0: low-grade vs. 1: high-grade, log-rank test p-value = 1.695^−10^, Figure 2.a1), hierarchical clusters (high vs. low, log-rank test p-value = 3.299^−12^, Figure 2.a2), risk group by median (high vs. low, log-rank test p-value = 9.651^−10^, Figure 2.a3). For Wilms tumor, the Kaplan-Meier curves of patient subtypes (Figure 2.b) were split by cancer type (FHWT vs. DAWT, log-rank test p-value= 0.4029, Figure 2.b1), hierarchical clusters (high vs. low, log-rank test p-value=0.3358, Figure 2.b2), risk group by median (high vs low, log-rank test p-value = 0.6587, Figure 2.b3). The results show that PCGS improves downstream tasks like patient subtyping, as reflected by stronger survival separation in the hierarchical clusters with smaller log-rank test p-values.

**Figure 2.**
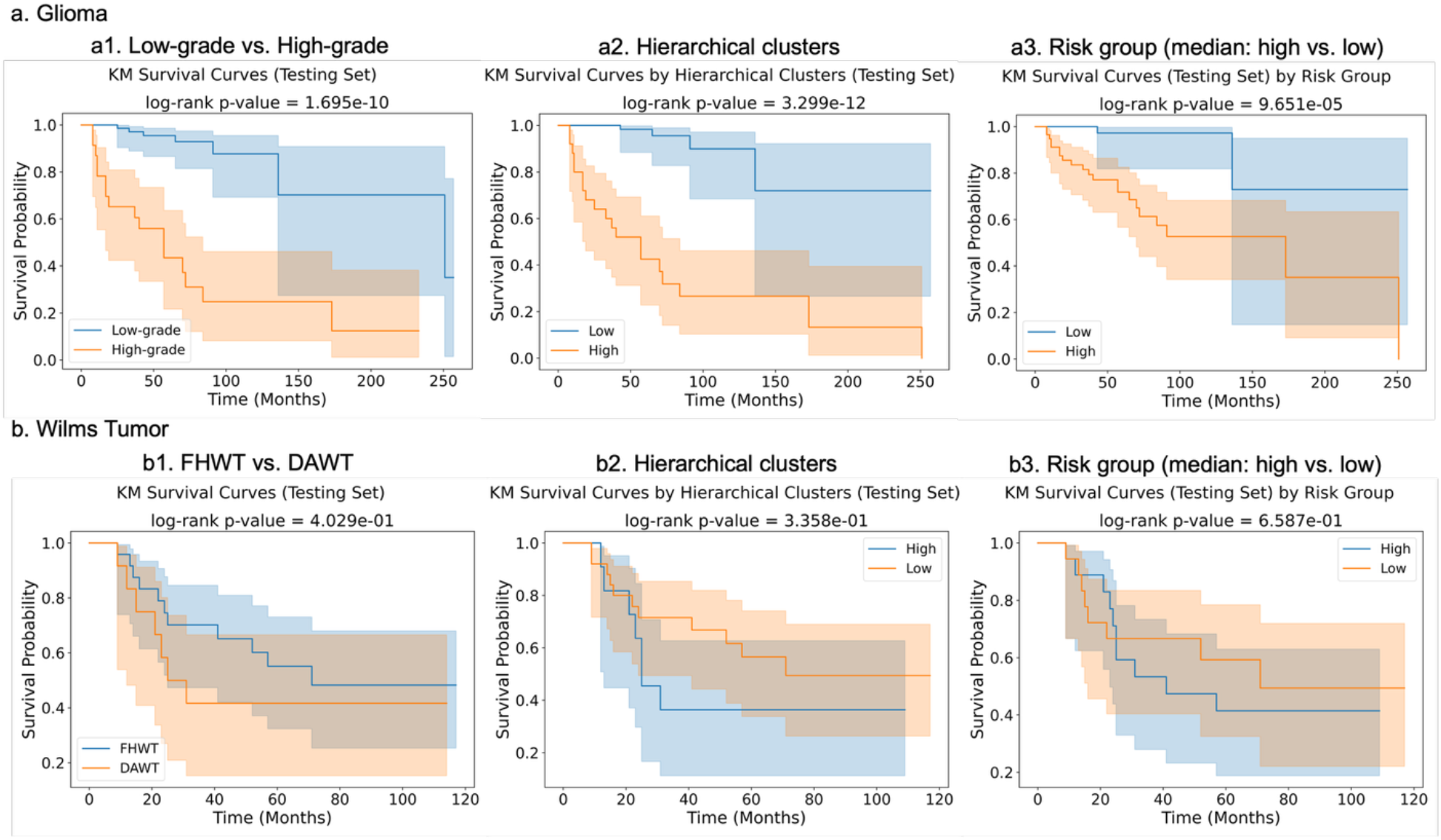
Survival analysis for patient subtypes based on test sets using GAT and cross-attention fusion. For glioma, the Kaplan-Meier curves of patient subtypes (Figure 2.a) were split by cancer type (0: low-grade vs. 1: high-grade, shown in Figure 2.a1), hierarchical clusters (Figure 2.a2), risk group by median (high vs. low, Figure 2.a3). For Wilms tumor, the Kaplan-Meier curves of patient subtypes (Figure 2.b) were split by cancer type (FHWT vs. DAWT, shown in Figure 2.b1), hierarchical clusters (Figure 2.b2), risk group by median (high vs low, Figure 2.b3).

Shapley value-based feature attribution enables GNN model explainability by estimating and ranking the importance scores for input features from each omics type, given specific background samples as baselines. Exemplary beeswarm plots (Figure 3) present the top genes ranked and selected by their mean Shapley values aggregated across samples to estimate the positive and negative influence of genes on the risk scores of pediatric glioma patients, using samples and gene expression (mRNA) data in the testing sets and GCN with cross-attention fusion layers. Randomly selected samples (Figure 3.a), samples from high-grade glioma groups (Figure 3.b), and samples from low-grade glioma groups (Figure 3.c) are selected as the baselines for estimating the contribution of mRNA biomarkers. For example, it suggests that the upregulation of CENPA will contribute to the increase of predicted risk scores (i.e., poor prognosis). As a protein involved in cell division and chromosome segregation, CENPA is upregulated in gliomas, especially glioblastoma, and is associated with poor prognosis, tumor aggressiveness, and immune infiltration ^34^. Other selected genes and their annotations, including expression status, impact on prognosis, and association with glioma grades, are consistent with previous publications (Table 3).

**Table 3.**
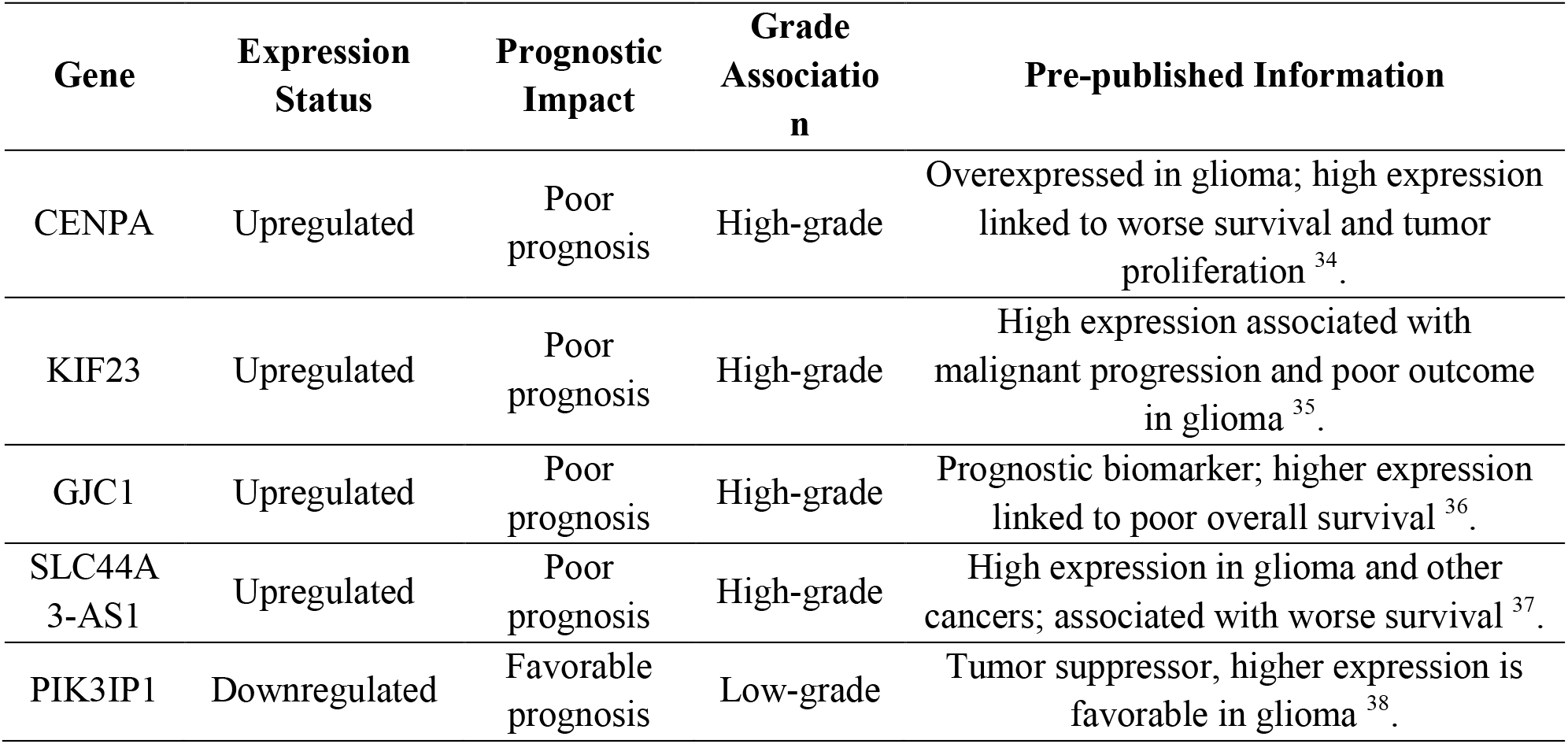
Selected top genes explaining survival for glioma.

**Figure 3.**
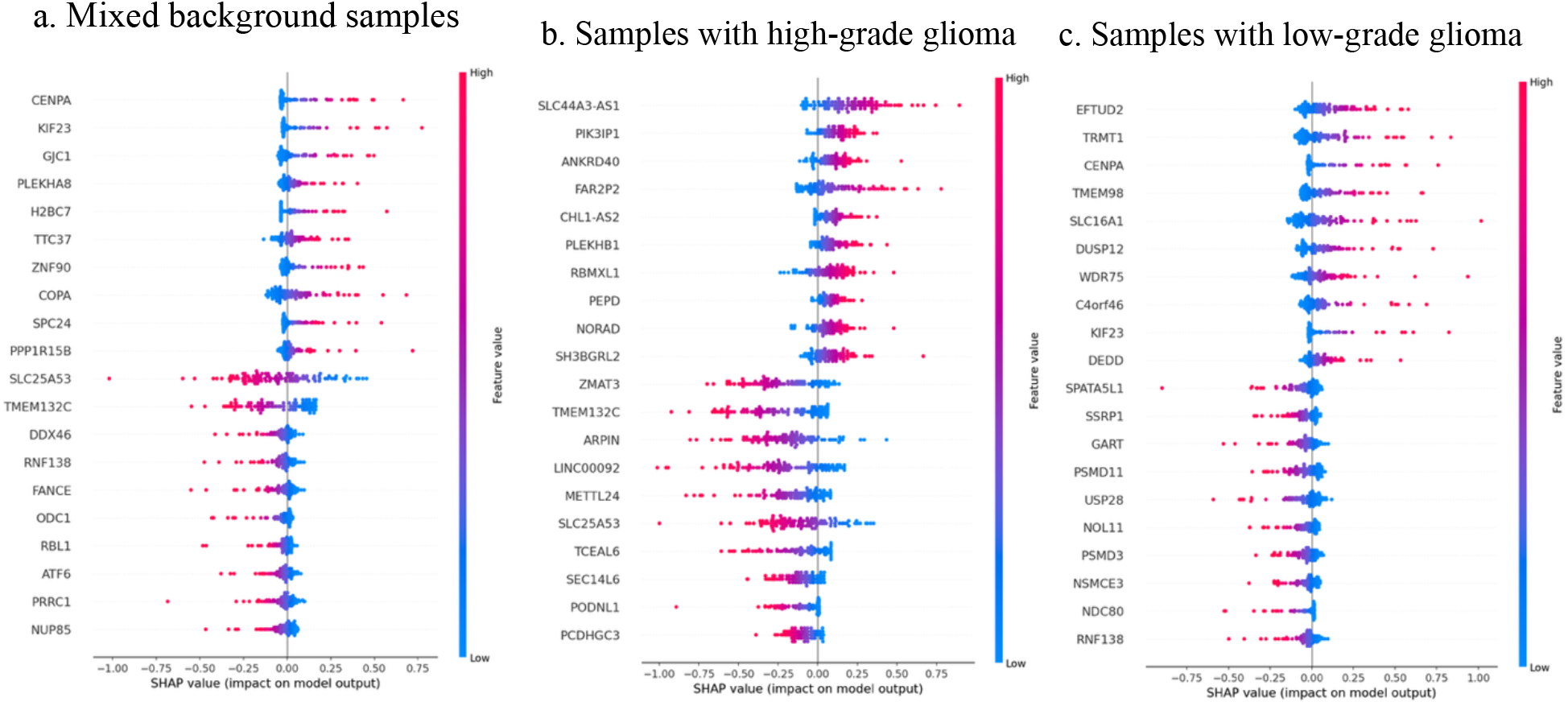
Shapley values of top genes from gene expression (mRNA) data of testing sets influencing glioma risk scores or survival based on GCN with cross-attention fusion. Genes were ranked and top 10 positive and negative ones respectively were selected by their mean Shapley values aggregated across samples in the testing sets. Randomly selected samples (Figure 3.a), samples from high-grade glioma groups (Figure 3.b), and samples from low-grade glioma groups (Figure 3.c) are used as the baselines for estimating Shapley values of mRNA biomarkers. In each instance, the explanation is visualized as a single dot per feature row, where the x-axis position reflects the SHAP value for that feature. Dots accumulate along each row to indicate density, while color represents the original feature value, ranging from low (blue) to high (red).

## 4. Discussion

This work, PCGS, offers a model framework for implementing explainable GNNs using Shapley values for feature attribution, enabling biomarker discovery, risk group identification, and survival analysis in pediatric cancers. Multi-omics GNN frameworks incorporating cross-attention and multi-head tasks, such as classification and survival analysis, can be employed to identify key biomarkers and stratify pediatric cancer patients to predict disease outcomes. SHAP-based methods further enable the estimation and comparison of feature importance to prioritize biomarkers across different baseline settings, enhancing the interpretability of classification and survival outcomes.

Due to the availability of processed omics data, only glioma and Wilms tumor cohorts were used to validate the model framework’s feasibility, performance, and explainability. Although the baseline models achieved comparable performance to the graph AI models in the glioma cohort, the integration of GNN and cross-attention fusion layers demonstrated greater performance in both classification and survival prediction tasks, particularly as evidenced by higher C-index values for risk-score prediction in the Wilms tumor cohort. Furthermore, our Shapley value-based explainable GNN model will not only be highly applicable on bulk genomics data via CCDI Data Ecosystem beyond glioma and Wilms tumor ^14^, but also it is extensible to be applied to single-cell spatial transcriptomics data, such as the Single-cell Pediatric Cancer Atlas Portal (ScPCA) ^39^.

Except for post-hoc Shapley value–based feature attribution for GNN explainability, an integrated Shapley value-based explanation model can be incorporated during training to compute importance scores for all edges connected to a target node, thereby improving the fidelity of the generated explanations. Thus, future efforts aimed at extensible implementation and the development of novel XAI methodologies will further advance the application of explainable GNN models for biomarker and drug discovery in pediatric oncology precision medicine.

To estimate whether the model performance will be affected by the number of pre-selected features, different numbers of pre-selected features were selected for glioma and Wilms tumor cohorts using thresholds in data preprocessing. Considering the difference of the number of raw features in the two cancer cohorts, 100, 200, 500, 1000, and 1500 features were preselected for glioma, while 100, 200, 300, 400, and 500 features were preselected for Wilms tumor. The model performance metrics such as accuracy, F1 score, and C-index values were estimated and averaged by 5-fold cross-validation for various model settings (Figure 4). As the number of preselected features increased, model performance remained largely unchanged across all configurations in the Wilms tumor cohort, whereas in the glioma cohort, performance initially improved for certain models while remaining stable for others. The tables of mean and standard deviation values of accuracy, F1 score, and C-index values for both glioma and Wilms tumor cohorts are presented in the supplementary materials.

**Figure 4.**
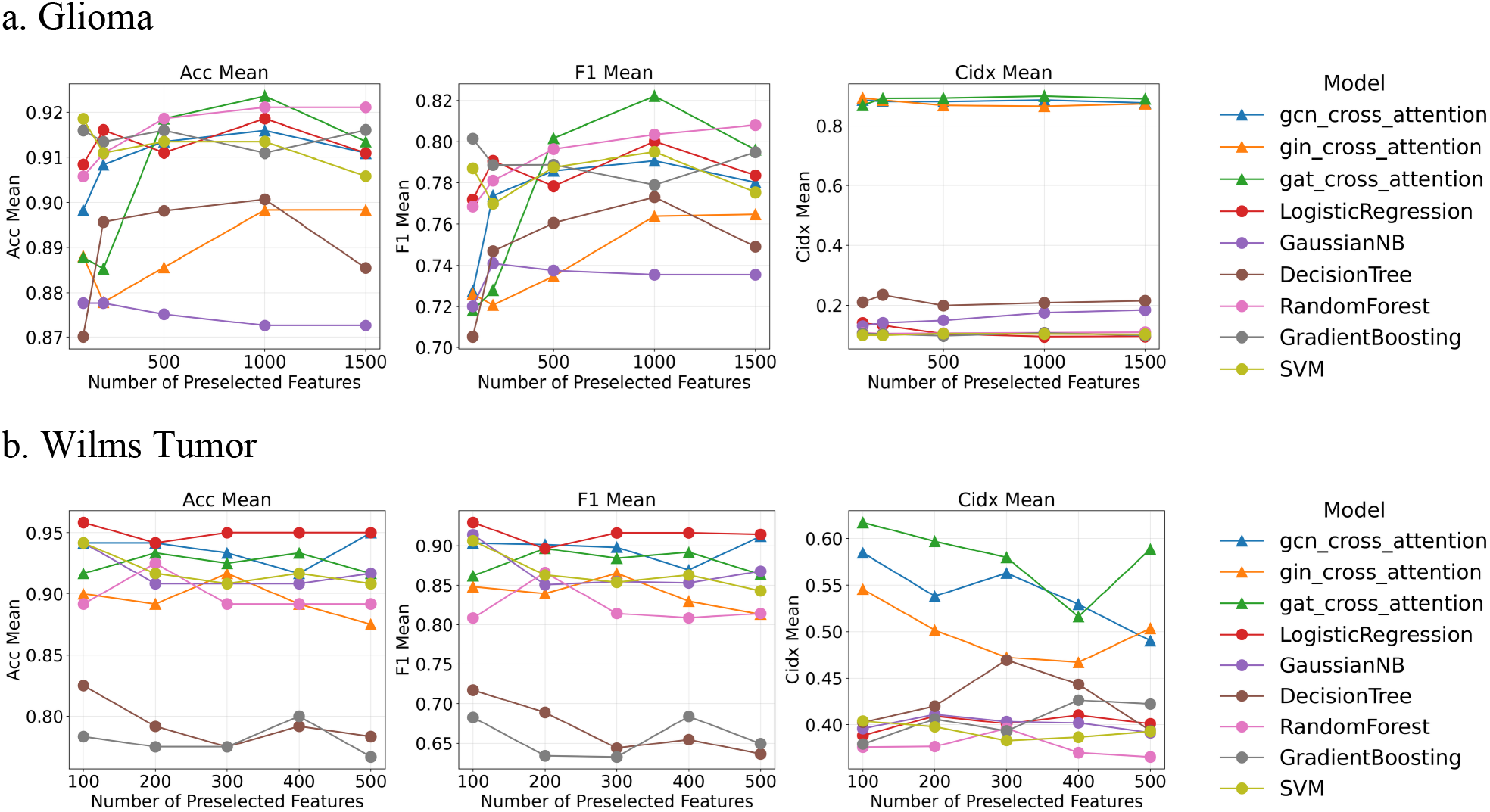
Influence of number of pre-selected feature number on model performance. 100, 200, 500, 1000, and 1500 features were preselected for glioma (Figure 4.a), while 100, 200, 300, 400, and 500 features were preselected for Wilms tumor (Figure 4.b). The mean values of model performance metrics such as accuracy, F1 score, and C-index values were estimated by 5-fold cross-validation and plotted under different preselected feature numbers for various model settings.

It should be noted that Shapley values-based feature attribution approach also has several limitations. First, post-hoc Shapley value estimation can alleviate the computational complexity for explainability^20^, but it may introduce variance or instability in estimated contributions. Second, the results are sensitive to the choice of background distribution or baseline input, which can influence the magnitude and direction of estimated contributions. Moreover, correlated or redundant features tend to share distributed importance, limiting interpretability in biological contexts. Furthermore, estimated Shapley values quantify feature contribution in a predictive sense but do not imply causation, so additional statistical analysis coupled with experimental validation are needed to identify true causal or biologically dominant predictors.

## Data Availability

All data produced are available online at:
https://www.cbioportal.org/
https://pedcbioportal.kidsfirstdrc.org/
https://clinicalcommons.ccdi.cancer.gov/home

https://pedcbioportal.kidsfirstdrc.org/

https://clinicalcommons.ccdi.cancer.gov/home

